# Inequalities in access to paid sick leave among workers in England and Wales

**DOI:** 10.1101/2022.01.30.22270112

**Authors:** Parth Patel, Sarah Beale, Vincent Nguyen, Isobel Braithwaite, Thomas E Byrne, Wing Lam Erica Fong, Ellen Fragaszy, Cyril Geismar, Susan Hoskins, Annalan M D Navaratnam, Madhumita Shrotri, Jana Kovar, Andrew C Hayward, Robert W Aldridge

## Abstract

**Background:** It is poorly understood which workers lack access to sick pay in England and Wales. This evidence gap has been of particular interest in the context of the Covid-19 epidemic given the relationship between presenteeism and infectious disease transmission.

**Method:** This cross-sectional analysis is nested within a large community cohort study of Covid-19 epidemiology in England and Wales (Virus Watch). An online survey in February 2021 asked participants if they had access to paid sick leave. We use a fixed effect logistic regression model to examine sociodemographic factors associated with lacking access to sick pay.

**Results:** 8,874 participants in work responded to the survey item about access to sick pay. Of those, 5,864 (66%) report having access to sick pay, 2,218 (25%) report no access to sick pay and 792 (8.9%) were unsure. Workers aged 45-64 (OR 1.72) and over 65 (OR 5.26) are more likely to lack access to sick pay compared to workers aged 25-44. South Asian workers (OR 1.40) and those from Other minority ethnic backgrounds (OR 2.93) are more likely to lack access to sick pay compared to White British workers. Workers in low income households (OR 1.43-2.53) and those with working class occupations (OR 2.04-5.29) are also more likely to lack access to sick pay compared to those in high income households and managerial occupations.

**Discussion:** Unwarranted age and race inequalities in sick pay access are suggestive of labour market discrimination. Occupational differences are also cause for concern. Policymakers should consider expanding access to sick pay to mitigate transmission of Covid-19 and other endemic infectious disease epidemics in the community.

## Introduction

An estimated two million employees in the UK do not earn enough to be eligible for statutory sick.^1,2^ For those who do, mandatory paid sick leave as a proportion of previous earnings is among the lowest of the countries constituting the Organisation for Economic and Co-operation and Development (OECD).^3^ Employees on casual or flexible contracts (including those on zero hour contracts) have a legal right to statutory sick pay if they are able prove their average earnings are above the eligibility threshold, which can be challenging. Statutory sick pay in the UK does not extend to those who are self-employed.

There has been considerable policy attention to statutory sick pay in the context of the Covid-19 pandemic.^2,3,14,15^ From a living standards perspective, limited access to paid sick leave risks income loss and destitution for low income households during periods of Covid-19 illness or self-isolation. From a public health perspective, the risk of income loss may encourage presenteeism and drive Covid-19 transmission in the community. For example, fear of loss income was a common reason given for non-adherence with self-isolation among workers during the 2003 SARS outbreak in Toronto.^4^ Furthermore, the introduction of mandatory sick pay policies across US states have been associated with lower infection disease rates in the community and mitigation of flu epidemics.^5,6^

Official UK government estimates on sick pay coverage come from a 2014 survey of 2,030 employees in Great Britain eligible for sick pay found that of employees eligible for sick pay, 26% receive the statutory minimum rate, 57% receive sick pay above this minimum and 17% don’t know.^7^ But this survey does not capture workers lacking access to paid sick leave. The characteristics of that group can only indirectly be inferred by examining workers earning below the income threshold to access statutory sick pay using data from the Office of National Statistics’ Labour Force Survey.^16^ This is imperfect and assumes all workers earnings above the eligibility threshold will automatically have access to paid sick leave. It is therefore limited in its ability to infer unwarranted labour market inequalities in access to paid sick leave.

Inequalities in access to sick pay among UK workers are poorly understood. In that context, this short communication exploits data from the Virus Watch study to examine factors associated with access to sick pay.

## Methods

### Study design and procedure

Data for this analysis were collected as part of the Virus Watch study, a large prospective household cohort study of Covid-19 transmission in England and Wales. The full study design and methodology has been described elsewhere.^8^ Participants were recruited into the Virus Watch study using a range of methods including by post, social media, SMS messages or personalised letters from General Practices with tokens of appreciation for participation. Participants were eligible if all household members agreed to take part, if they had access to the internet (Wi-Fi, fixed or mobile network) and an email address. At least one household member had to be able to read English to complete the surveys. Participants were not eligible if their household was larger than 6 people (due to limitations of the online survey infrastructure).

After enrolling in the study, an initial baseline survey collected demographic, occupation, social and medical history data from participants. Thereafter, participants were surveyed weekly (contacted by email) on the presence or absence of symptoms that could indicate COVID-19 disease, activities undertaken during that period, SARS-CoV-2 test results and COVID-19 vaccine uptake in the previous week. Bespoke monthly surveys collected detailed information on potential determinants of COVID-19 illness. A February 2021 survey focussed on financial and work-related determinants of COVID-19 illness included an item relating to sick pay access. The analysis in this communication is limited to those who responded to this February 2021 survey and have recorded employment or self-employment status.

### Exposures

The exposures of interest were demographic and social variables that could be associated with differing sick pay access among working adults. These were age; sex; region; ethnicity; household income; and occupation. Data on exposure variables were collected through the baseline survey completed on entry into the Virus Watch study.

### Outcome

The outcome of interest was self-reported access to sick pay. Participants were able to report ‘Yes’, ‘No’, ‘Unsure’ and ‘Not applicable’ when asked if they had access to paid sick leave if required. Those who responded ‘Not applicable’ were excluded from this study. Outcome data was collected between 17–28 February 2021.

### Statistical analysis

To model the association between covariates and access to sick pay, we conducted univariable and multivariable fixed effects logistic regression models in R 4.0.3 to derive relative odds ratios and 95% confidence intervals. A sensitivity analysis controlling for self-employment status (employed vs self-employed) was also conducted, as it was felt this may confound the relationship between ethnicity and sick pay access.

### Patient and public involvement

The study team worked with the Race Equality Foundation and Doctors of the World who advised on the inclusion of people from minority ethnic backgrounds in Virus Watch and set up a community advisory group to inform the ongoing design and dissemination of health equity aspects of Virus Watch. This advisory group, consisting of lay members of the public, community leaders, charities and policy experts, suggested the analysis described in this article.

## Results

Table 1 reports the characteristics of Virus Watch participants who are in work and responded to the 17 February 2021 survey question inquiring about access to paid sick leave (n=8,874). Of those, 5,864 (66%) report having access to paid sick leave, 2,218 (25%) report no access to paid sick leave and 792 (8.9%) were unsure. Table 2 describes characteristics of those with and without access to sick pay.

**Table 1.**
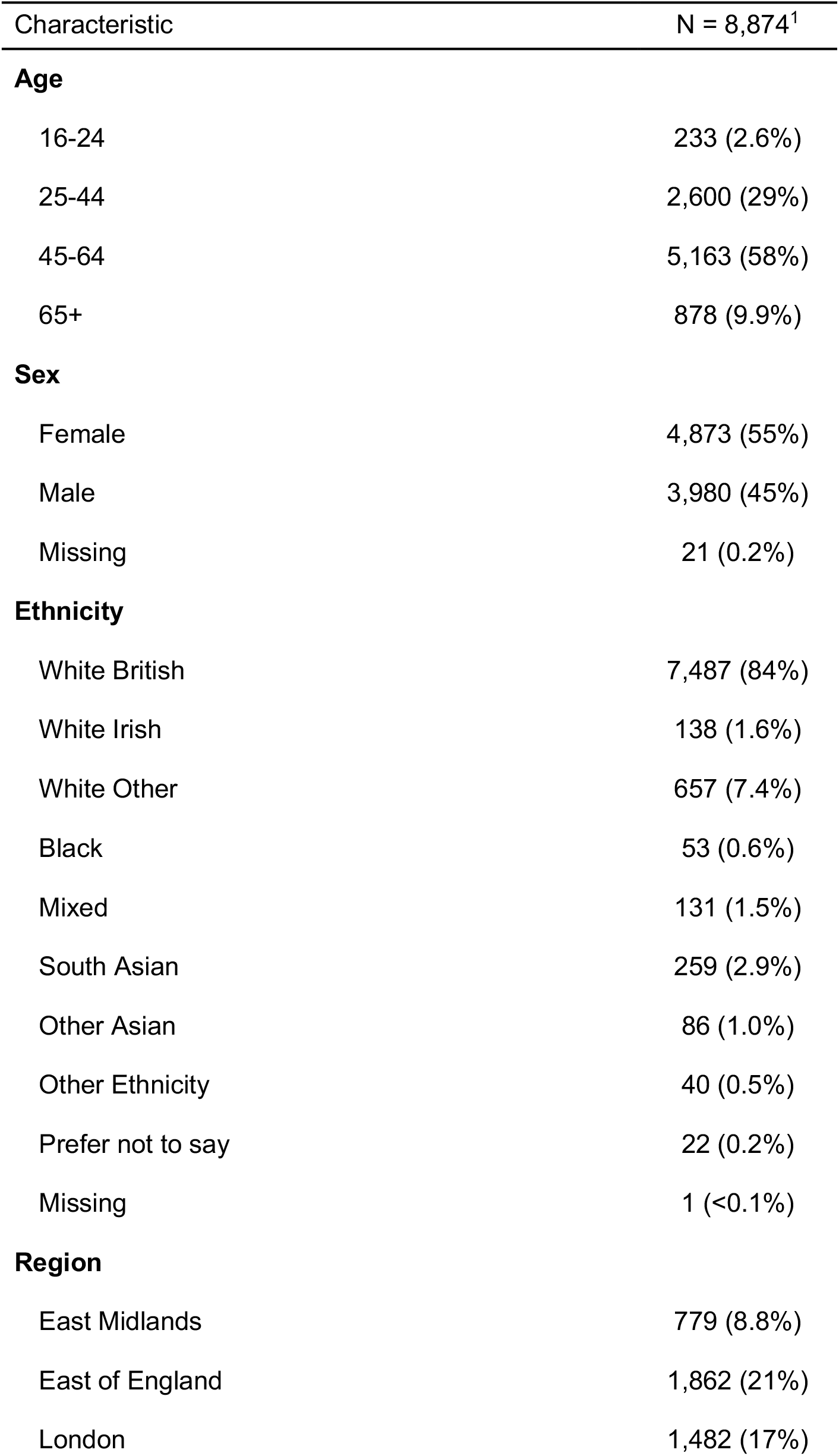
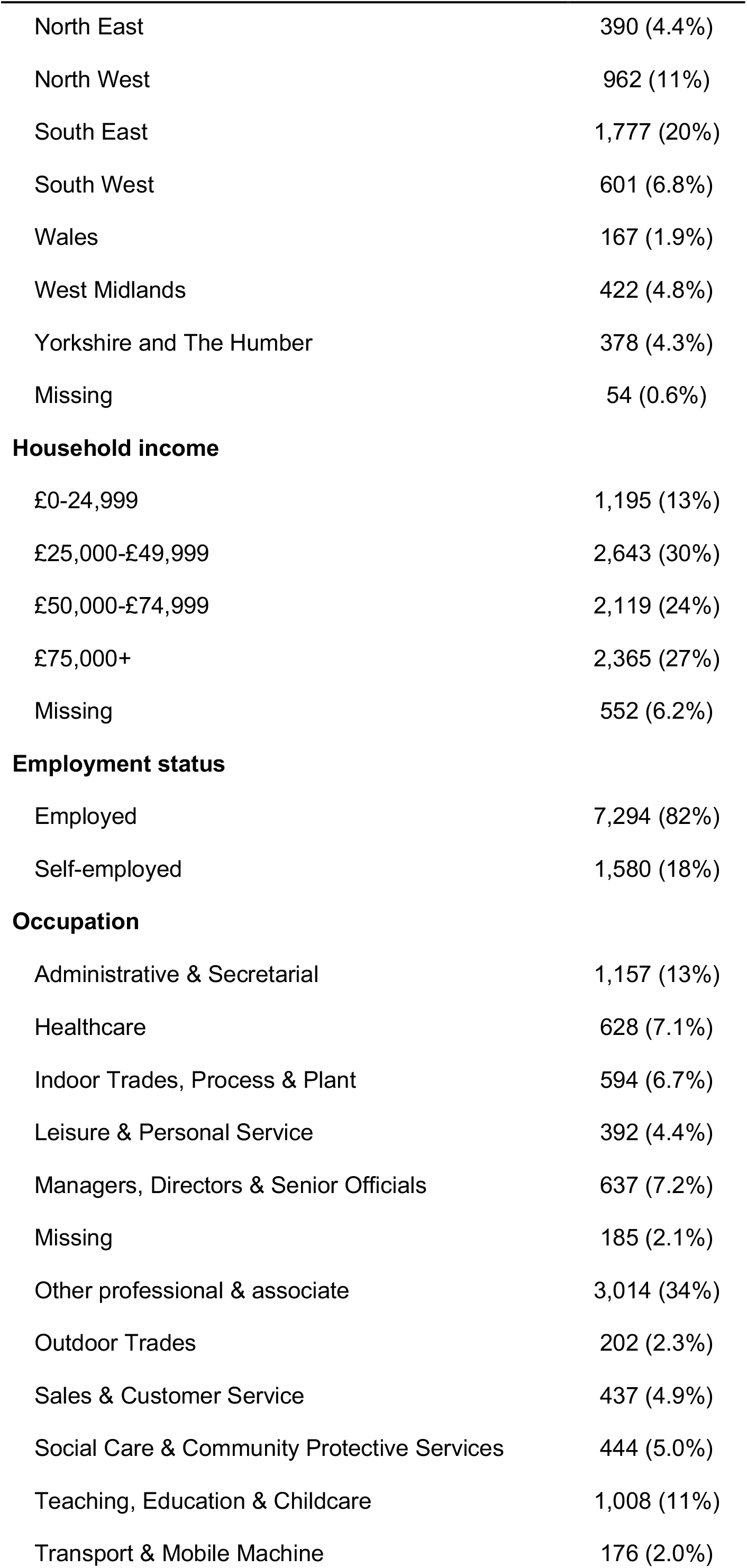
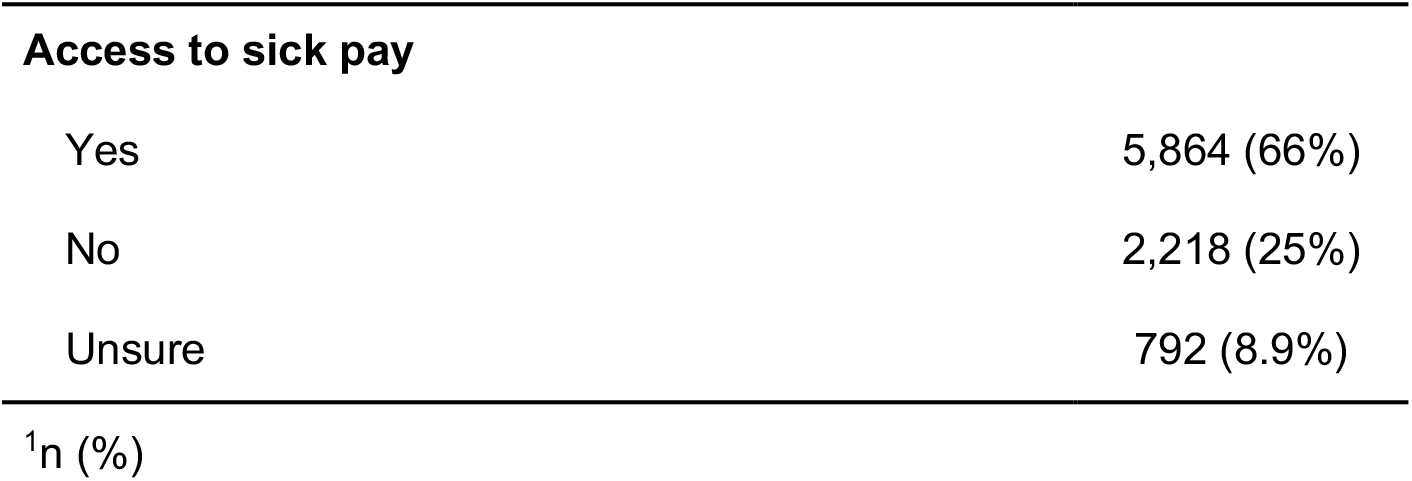
Sociodemographic characteristics of survey respondents.

| Characteristic | N = 8,874<sup>1</sup> |
| --- | --- |
| **Age** |  |
| 16-24 | 233 (2.6%) |
| 25-44 | 2,600 (29%) |
| 45-64 | 5,163 (58%) |
| 65+ | 878 (9.9%) |
| **Sex** |  |
| Female | 4,873 (55%) |
| Male | 3,980 (45%) |
| Missing | 21 (0.2%) |
| **Ethnicity** |  |
| White British | 7,487 (84%) |
| White Irish | 138 (1.6%) |
| White Other | 657 (7.4%) |
| Black | 53 (0.6%) |
| Mixed | 131 (1.5%) |
| South Asian | 259 (2.9%) |
| Other Asian | 86 (1.0%) |
| Other Ethnicity | 40 (0.5%) |
| Prefer not to say | 22 (0.2%) |
| Missing | 1 (<0.1%) |
| **Region** |  |
| East Midlands | 779 (8.8%) |
| East of England | 1,862 (21%) |
| London | 1,482 (17%) |
| North East | 390 (4.4%) |
| North West | 962 (11%) |
| South East | 1,777 (20%) |
| South West | 601 (6.8%) |
| Wales | 167 (1.9%) |
| West Midlands | 422 (4.8%) |
| Yorkshire and The Humber | 378 (4.3%) |
| Missing | 54 (0.6%) |
| **Household income** |  |
| £0-24,999 | 1,195 (13%) |
| £25,000-£49,999 | 2,643 (30%) |
| £50,000-£74,999 | 2,119 (24%) |
| £75,000+ | 2,365 (27%) |
| Missing | 552 (6.2%) |
| **Employment status** |  |
| Employed | 7,294 (82%) |
| Self-employed | 1,580 (18%) |
| **Occupation** |  |
| Administrative & Secretarial | 1,157 (13%) |
| Healthcare | 628 (7.1%) |
| Indoor Trades, Process & Plant | 594 (6.7%) |
| Leisure & Personal Service | 392 (4.4%) |
| Managers, Directors & Senior Officials | 637 (7.2%) |
| Missing | 185 (2.1%) |
| Other professional & associate | 3,014 (34%) |
| Outdoor Trades | 202 (2.3%) |
| Sales & Customer Service | 437 (4.9%) |
| Social Care & Community Protective Services | 444 (5.0%) |
| Teaching, Education & Childcare | 1,008 (11%) |
| Transport & Mobile Machine | 176 (2.0%) |
| **Access to sick pay** |  |
| Yes | 5,864 (66%) |
| No | 2,218 (25%) |
| Unsure | 792 (8.9%) |
| <sup>1</sup>n (%) |  |

**Table 2.** Description of participant characteristics by self-reported access to sick pay.

| Characteristic | Access to sick pay |  |  |
| --- | --- | --- | --- |
|  | Yes, N = 5,864<sup>1</sup> | No, N = 2,218<sup>1</sup> | Unsure, N = 792<sup>1</sup> |
| **Age** |  |  |  |
| 16-24 | 161 (69%) | 39 (17%) | 33 (14%) |
| 25-44 | 2,001 (77%) | 408 (16%) | 191 (7.3%) |
| 45-64 | 3,382 (66%) | 1,368 (26%) | 413 (8.0%) |
| 65+ | 320 (36%) | 403 (46%) | 155 (18%) |
| **Sex** |  |  |  |
| Female | 3,358 (69%) | 1,098 (23%) | 417 (8.6%) |
| Male | 2,491 (63%) | 1,115 (28%) | 374 (9.4%) |
| Missing | 15 (71%) | 5 (24%) | 1 (4.8%) |
| **Ethnicity** |  |  |  |
| White British | 4,935 (66%) | 1,924 (26%) | 628 (8.4%) |
| White Irish | 88 (64%) | 35 (25%) | 15 (11%) |
| White Other | 457 (70%) | 128 (19%) | 72 (11%) |
| Black | 35 (66%) | 10 (19%) | 8 (15%) |
| Mixed | 92 (70%) | 30 (23%) | 9 (6.9%) |
| South Asian | 168 (65%) | 53 (20%) | 38 (15%) |
| Other Asian | 55 (64%) | 17 (20%) | 14 (16%) |
| Other Ethnicity | 19 (48%) | 15 (38%) | 6 (15%) |
| Prefer not to say | 14 (64%) | 6 (27%) | 2 (9.1%) |
| Missing | 1 (100%) | 0 (0%) | 0 (0%) |
| **Region** |  |  |  |
|  | Yes, N = 5,864<sup>1</sup> | No, N = 2,218<sup>1</sup> | Unsure, N = 792<sup>1</sup> |
| East Midlands | 518 (66%) | 202 (26%) | 59 (7.6%) |
| East of England | 1,200 (64%) | 509 (27%) | 153 (8.2%) |
| London | 994 (67%) | 336 (23%) | 152 (10%) |
| North East | 280 (72%) | 79 (20%) | 31 (7.9%) |
| North West | 672 (70%) | 218 (23%) | 72 (7.5%) |
| South East | 1,113 (63%) | 480 (27%) | 184 (10%) |
| South West | 399 (66%) | 162 (27%) | 40 (6.7%) |
| Wales | 113 (68%) | 38 (23%) | 16 (9.6%) |
| West Midlands | 282 (67%) | 100 (24%) | 40 (9.5%) |
| Yorkshire and The Humber | 256 (68%) | 86 (23%) | 36 (9.5%) |
| Missing | 37 (69%) | 8 (15%) | 9 (17%) |
| **Household income** |  |  |  |
| £0-24,999 | 586 (49%) | 409 (34%) | 200 (17%) |
| £25,000-£49,999 | 1,676 (63%) | 745 (28%) | 222 (8.4%) |
| £50,000-£74,999 | 1,525 (72%) | 457 (22%) | 137 (6.5%) |
| £75,000+ | 1,757 (74%) | 440 (19%) | 168 (7.1%) |
| Missing | 320 (58%) | 167 (30%) | 65 (12%) |
| **Employment status** |  |  |  |
| Employed | 5,773 (79%) | 954 (13%) | 567 (7.8%) |
| Self-employed | 91 (5.8%) | 1,264 (80%) | 225 (14%) |
| **Occupation** |  |  |  |
| Administrative & Secretarial | 860 (74%) | 200 (17%) | 97 (8.4%) |
| Healthcare | 422 (67%) | 150 (24%) | 56 (8.9%) |
| Indoor Trades, Process & Plant | 311 (52%) | 222 (37%) | 61 (10%) |
| Leisure & Personal Service | 176 (45%) | 162 (41%) | 54 (14%) |
| Managers, Directors & Senior Officials | 462 (73%) | 118 (19%) | 57 (8.9%) |
| Missing | 118 (64%) | 50 (27%) | 17 (9.2%) |
| Other professional & associate | 2,020 (67%) | 743 (25%) | 251 (8.3%) |
| Outdoor Trades | 55 (27%) | 121 (60%) | 26 (13%) |
|  | Yes, N = 5,864<sup>1</sup> | No, N = 2,218<sup>1</sup> | Unsure, N = 792<sup>1</sup> |
| Sales & Customer Service | 278 (64%) | 116 (27%) | 43 (9.8%) |
| Social Care & Community Protective Services | 307 (69%) | 104 (23%) | 33 (7.4%) |
| Teaching, Education & Childcare | 772 (77%) | 163 (16%) | 73 (7.2%) |
| Transport & Mobile Machine | 83 (47%) | 69 (39%) | 24 (14%) |
|  |  |  | <sup>1</sup>n (%) |

Results of the univariable and multivariable logistic regression model including age, sex, region, ethnicity, household income and occupation are in Table 3. In the multivariable model, workers over the age of 65 (OR 5.26, 95CI 4.42-6.26) and between ages of 45 and 64 (OR 1.72, 95CI 1.53-1.93) have greater odds of lacking access to sick pay in reference to workers aged 25-44. South Asian workers (OR 1.40, 95CI 1.06-1.83) and ‘Other minority ethnic’ workers (OR 2.93, 95CI 1.54-5.59) also have greater odds of lacking access to sick pay compared to White British workers. It is worth noting that although not always statistically significant, workers from most minority ethnic backgrounds had elevated odds of lacking access to sick pay compared to White British workers. People in low income households are more likely to lack access to sick pay compared to those high income households, with households earning under £25,000 (OR 2.53, 95CI 2.15-2.98) and households earning £25,000-£49,999 (OR 1.43, 95CI 1.25-1.63) at greater odds of lacking access to sick pay if required than those in households earning above £75,000. Workers in leisure and personal service (OR 2.43, 95CI 1.84-3.21), indoor trades, process and plant (OR 2.03, 95CI 1.58-2.61), outdoor trades (OR 5.29, 95CI 3.67-7.72) and transport and mobile machinery (OR 2.04, 95CI 1.42-2.94) occupations are all more likely to lack access to sick pay compared to managers, directors and senior officials.

**Table 3.**
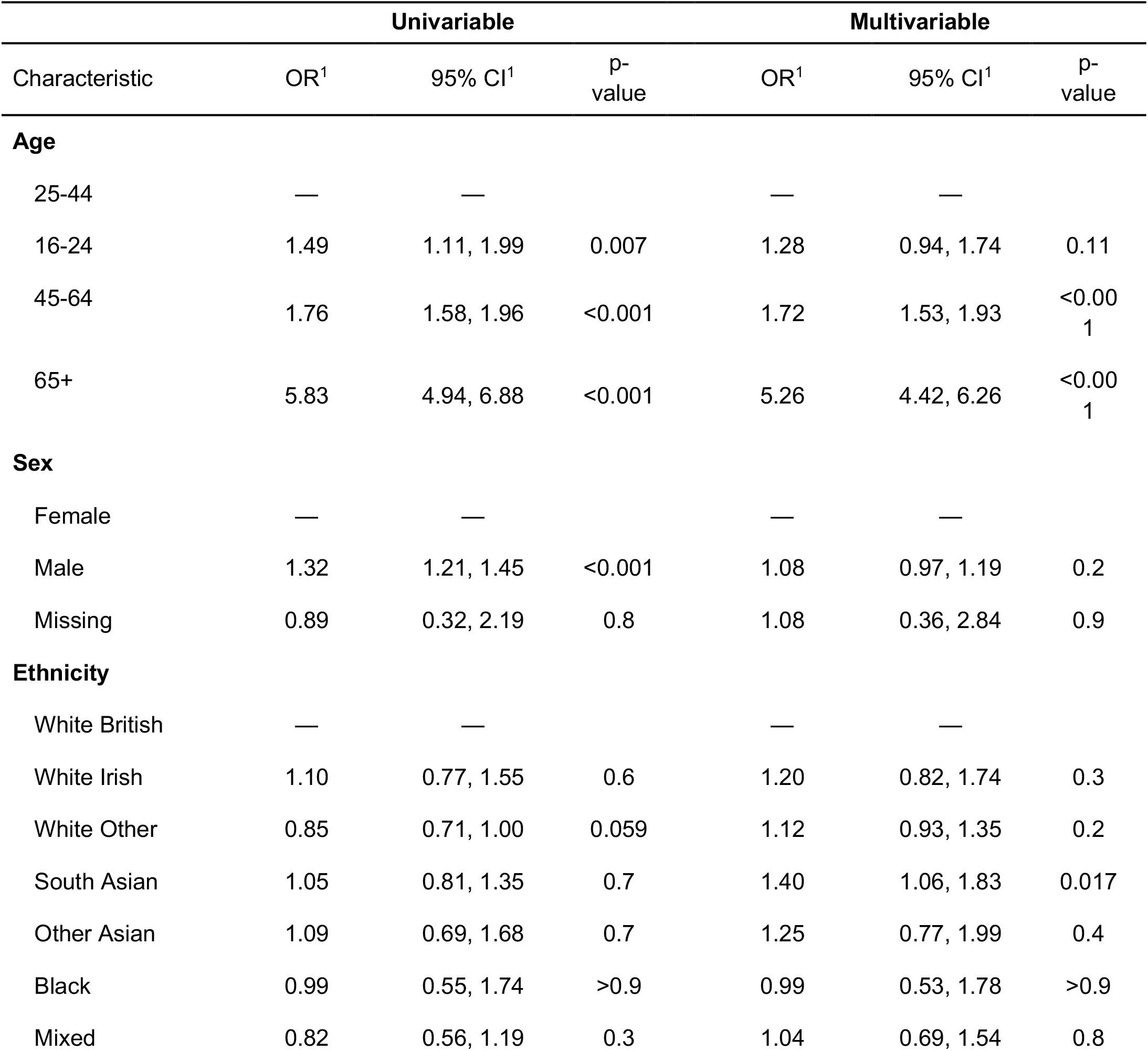
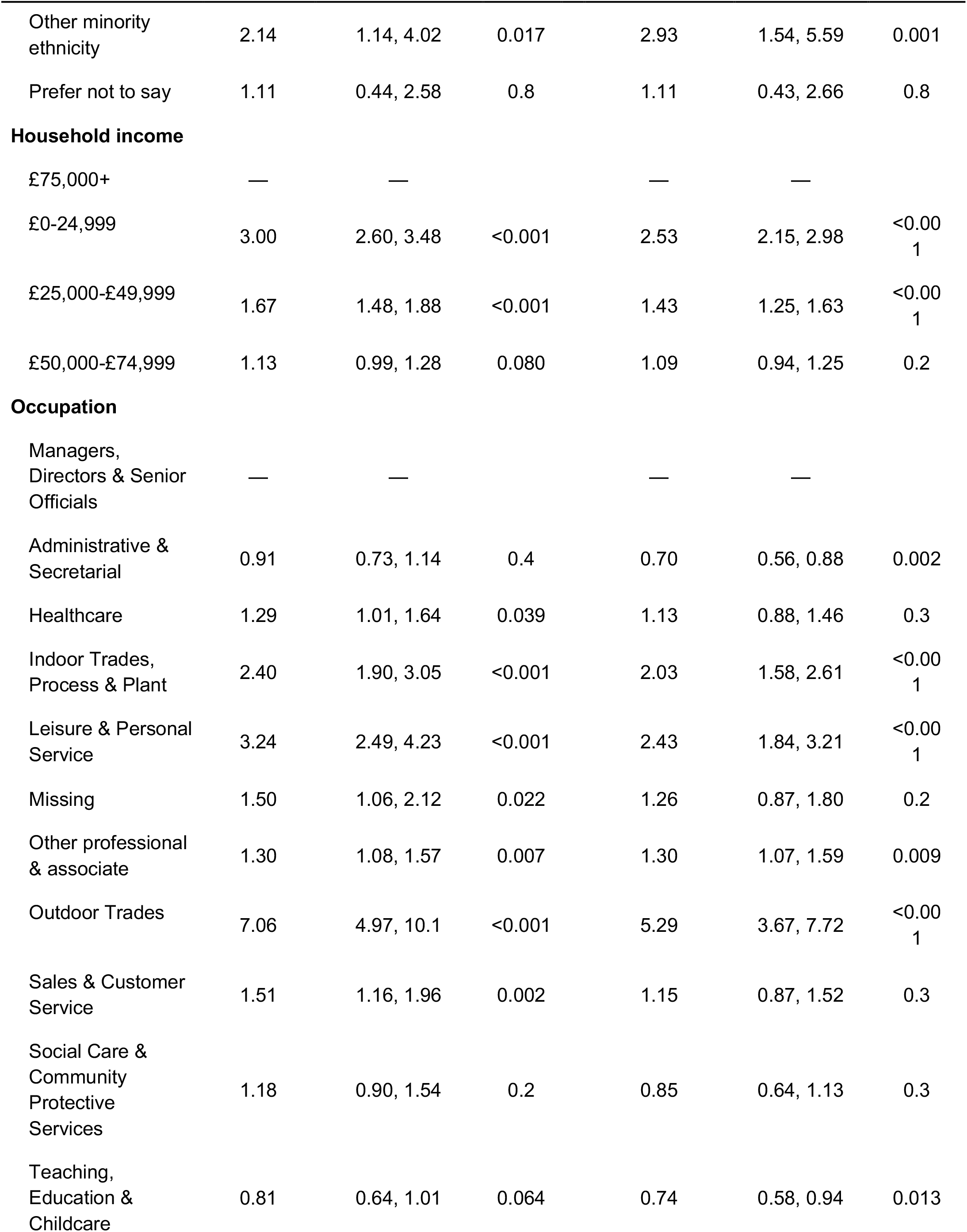
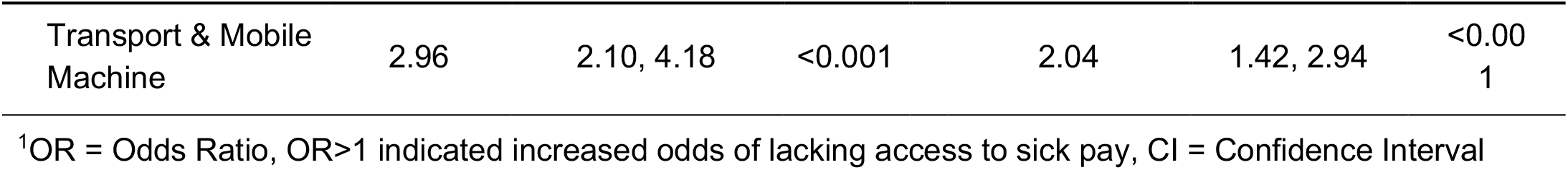
Univariable and multivariable logistic regression models examining the relationship between sociodemographic characteristics and lacking access to sick pay.

A sensitivity analysis controlling for self-employment status was consistent with these findings (Table 4). All observed differences in sick pay access between age, ethnic and income groups persisted. Indeed, ethnic contracts in access to sick pay heightened. 16-24 year old workers (OR 1.73, 95CI 1.25-2.38) are also at greater odds of lacking access to sick pay compared to 24-44 year old workers after adjusting for self-employment status. With the exception of outdoor trade occupations, all observed occupational differences in sick pay access persisted.

**Table 4.**
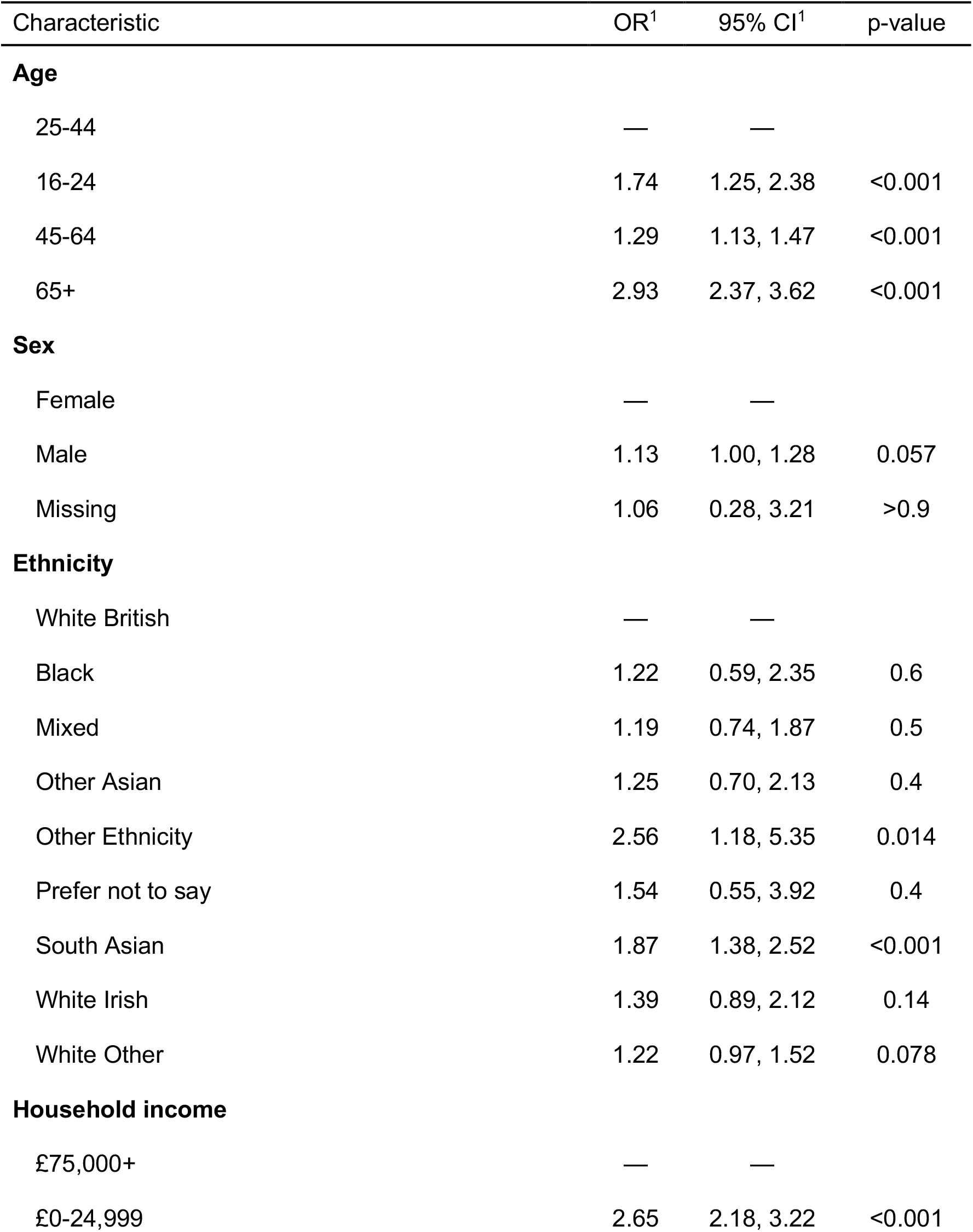
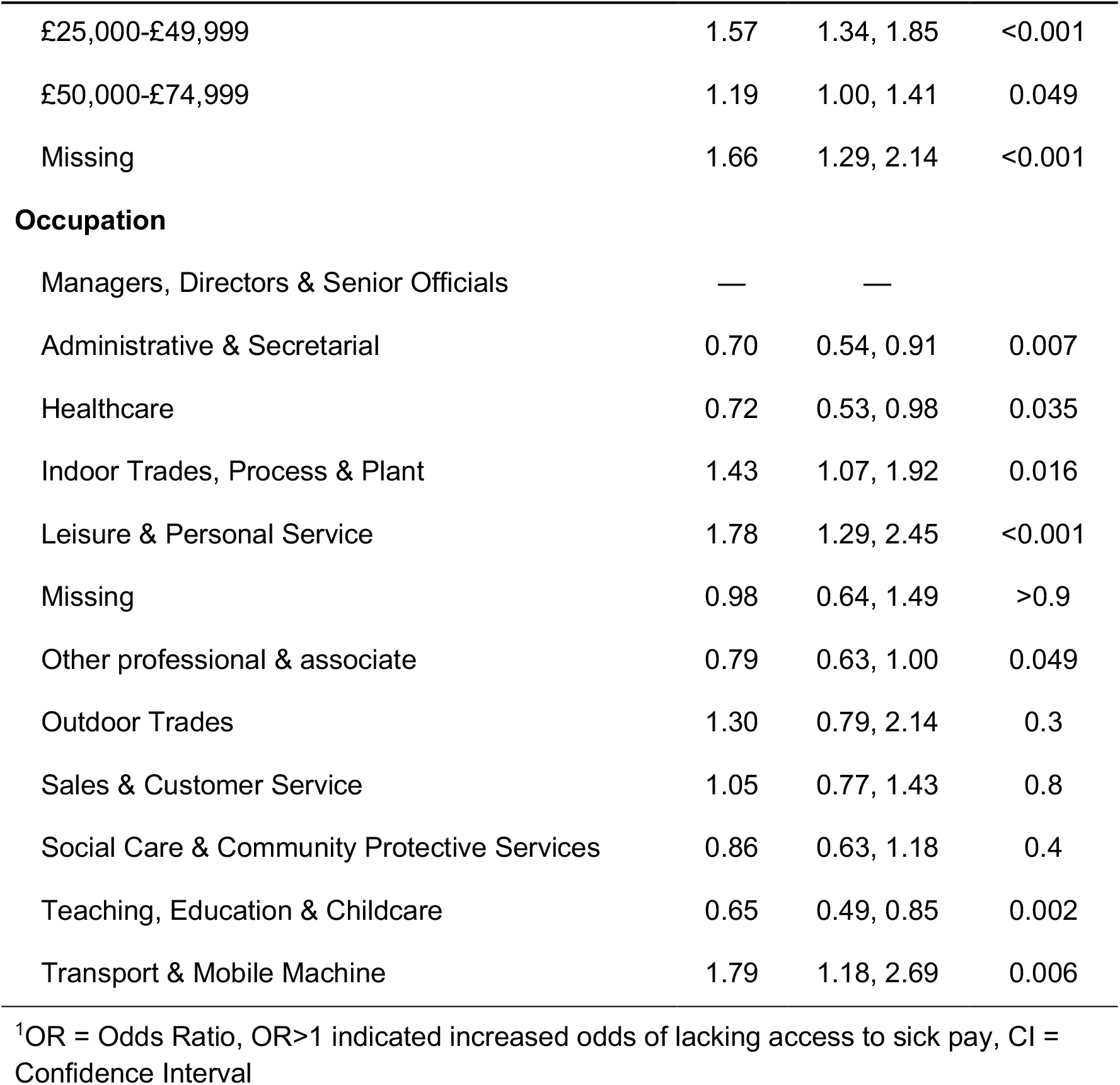
Sensitivity analysis multivariable logistic regression model including self-employment status as a covariate.

| Characteristic | OR<sup>1</sup> | 95% CI<sup>1</sup> | p-value |
| --- | --- | --- | --- |
| **Age** |  |  |  |
| 25-44 | — | — |  |
| 16-24 | 1.74 | 1.25, 2.38 | <0.001 |
| 45-64 | 1.29 | 1.13, 1.47 | <0.001 |
| 65+ | 2.93 | 2.37, 3.62 | <0.001 |
| **Sex** |  |  |  |
| Female | — | — |  |
| Male | 1.13 | 1.00, 1.28 | 0.057 |
| Missing | 1.06 | 0.28, 3.21 | >0.9 |
| **Ethnicity** |  |  |  |
| White British | — | — |  |
| Black | 1.22 | 0.59, 2.35 | 0.6 |
| Mixed | 1.19 | 0.74, 1.87 | 0.5 |
| Other Asian | 1.25 | 0.70, 2.13 | 0.4 |
| Other Ethnicity | 2.56 | 1.18, 5.35 | 0.014 |
| Prefer not to say | 1.54 | 0.55, 3.92 | 0.4 |
| South Asian | 1.87 | 1.38, 2.52 | <0.001 |
| White Irish | 1.39 | 0.89, 2.12 | 0.14 |
| White Other | 1.22 | 0.97, 1.52 | 0.078 |
| **Household income** |  |  |  |
| £75,000+ | — | — |  |
| £0-24,999 | 2.65 | 2.18, 3.22 | <0.001 |
| £25,000-£49,999 | 1.57 | 1.34, 1.85 | <0.001 |
| £50,000-£74,999 | 1.19 | 1.00, 1.41 | 0.049 |
| Missing | 1.66 | 1.29, 2.14 | <0.001 |
| **Occupation** |  |  |  |
| Managers, Directors & Senior Officials | — | — |  |
| Administrative & Secretarial | 0.70 | 0.54, 0.91 | 0.007 |
| Healthcare | 0.72 | 0.53, 0.98 | 0.035 |
| Indoor Trades, Process & Plant | 1.43 | 1.07, 1.92 | 0.016 |
| Leisure & Personal Service | 1.78 | 1.29, 2.45 | <0.001 |
| Missing | 0.98 | 0.64, 1.49 | >0.9 |
| Other professional & associate | 0.79 | 0.63, 1.00 | 0.049 |
| Outdoor Trades | 1.30 | 0.79, 2.14 | 0.3 |
| Sales & Customer Service | 1.05 | 0.77, 1.43 | 0.8 |
| Social Care & Community Protective Services | 0.86 | 0.63, 1.18 | 0.4 |
| Teaching, Education & Childcare | 0.65 | 0.49, 0.85 | 0.002 |
| Transport & Mobile Machine | 1.79 | 1.18, 2.69 | 0.006 |

<sup>1</sup>OR = Odds Ratio, OR>1 indicated increased odds of lacking access to sick pay, CI = Confidence Interval

## Discussion

Our findings reveal stark inequalities in access to paid sick leave among workers in England and Wales. Older workers, people of certain minority ethnic groups, workers in low-income households and those in working class occupations are more likely to lack access to paid sick leave. These differences were not explainable by self-employment status.

This cross-sectional analysis is nested in the larger Virus Watch prospective cohort study. By harnessing data already collected by this large cohort study of Covid-19 epidemiology, this analysis is able to fill an important policy-relevant evidence gap on inequalities in sick pay access without needing to duplicate efforts to generate research data. Individuals in the Virus Watch study are well distributed across England and Wales and the cohort is diverse in terms of age, sex, ethnicity, and socioeconomic composition.^8^ Given participation in the Virus Watch study is voluntary and sampling non-random, the cohort likely oversamples people concerned with COVID-19 and participants were more likely to be White British, over the age of 65 and have a higher income than the general population. Our multivariable regression analysis adjusted for age, ethnicity and income will reduce this sampling bias, but any residual confounding is likely to mean our findings overestimate the true magnitude of age disparities in sick pay access and underestimate ethnic and income disparities in sick pay access.

There is little public data or previous literature on sick pay coverage in the UK. A descriptive analysis of 3,974 UK workers found leisure and personal services, outdoor trades, transport and mobile machinery and indoor trades occupations have the lowest rates of paid sick leave coverage, in keeping with the findings of our study.^9^ A 2014 survey of 2,030 employees by the Department of Work and Pensions is limited employees eligible to access sick pay.^7^ It reported descriptive differences in the amount of sick pay workers receive, with a greater proportion of older workers and those in leisure and personal service occupations reporting access to only the minimum statutory rate. There has been greater study of disparities in sick leave access in the US, where ethnic inequalities have been documented.^10,11^

It is unsurprising that those in low income households are more likely to lack access to sick pay than those in high income households given statutory sick pay entitlement in the UK is conditional on earning above an income threshold. Inequalities in sick pay access between age and ethnic groups that cannot be explained by differences in income, occupation and employment status are suggestive of age- and race-based discrimination in the labour market. Occupations with elevated odds of lacking access to sick pay relative to managerial occupations are classifiable as manual occupations and unskilled non-manual occupations according to the widely used Goldthorpe class scheme.^17^ These occupations are referred to as *Working Class* in the Goldthorpe occupation-based class taxonomy.^12^ That working class occupations are most likely to lack access to sick pay is cause for concern with regard to inequalities in the labour market.

In the context of the Covid-19 pandemic, fear of income loss is likely to encourage presenteeism and SARS-CoV-2 transmission. A recent study of care homes in England found lower rates of SARS-CoV-2 transmission from staff when they had access to sick pay, compared to care homes where staff lacked access to statutory sick pay.^13^ Future research should investigate the relationship between access to sick pay and presenteeism in the community setting during the Covid-19 pandemic.

More broadly, paid sick leave has been judged an effective intervention to reduce transmission of SARS-CoV-2 across OECD countries.^3^ The UK government made some changes to statutory sick pay when the pandemic began by allowing eligible employees to receive sick pay during periods of self-isolation in addition to confirmed Covid-19 illness, and to receive sick pay from the first day of illness or self-isolation, rather than from the fourth day of illness as is the case for other illnesses. However, unlike around half of other OECD countries, the UK has not altered the replacement rate, nor has it modified the eligibility criteria to expand access to statutory sick pay. Given the inequalities we highlight in this paper, improving access to sick pay should be both an employment and health policy priority as Covid-19 becomes an endemic disease.

## Data Availability

All data produced in the present work are contained in the manuscript.

